# Rapid genomic surveillance of SARS-CoV-2 in a dense urban community using environmental (sewage) samples

**DOI:** 10.1101/2021.03.29.21254053

**Authors:** Rajindra Napit, Prajwol Manandhar, Ashok Chaudhary, Bishwo Shrestha, Ajit Poudel, Roji Raut, Saman Pradhan, Samita Raut, Sujala Mathema, Rajesh Rajbhandari, Sameer Dixit, Jessica S. Schwind, Christine K Johnson, Jonna K Mazet, Dibesh Karmacharya

## Abstract

Understanding disease burden and transmission dynamics in resource-limited, developing countries like Nepal is often challenging due to a lack of adequate surveillance systems. These issues are exacerbated by limited access to diagnostic and research facilities throughout the country. Nepal has one of the highest COVID-19 case rates (915 cases per 100,000 people) in South Asia, with densely-populated Kathmandu experiencing the highest number of cases. Swiftly identifying case clusters and introducing effective intervention programs is crucial to mounting an effective containment strategy. The rapid identification of circulating SARS-CoV-2 variants can also provide important information on viral evolution and epidemiology. Genomic-based environmental surveillance can help in the early detection of outbreaks before clinical cases are recognized, and identify viral micro-diversity that can be used for designing real-time risk-based interventions. This research aimed to develop a genomic-based environmental surveillance system by detecting and characterizing SARS-CoV-2 in sewage samples of Kathmandu using portable next-generation DNA sequencing devices. Out of 20 selected sites in the Kathmandu Valley, sewage samples from 16 (80%) sites had detectable SARS-CoV-2. A heat-map was created to visualize transmission activity in the community based on viral load intensity and corresponding geospatial data. Further, 41 mutations were observed in the SARS-CoV-2 genome. Some detected mutations (n=9, 2%) were novel and yet to be reported in the global database, with one indicating a frameshift deletion in the spike gene. We also observed more transition than transversion on detected mutations, indicating rapid viral evolution in the host. Our study has demonstrated the feasibility of rapidly obtaining vital information on community transmission and disease dynamics of SARS-CoV-2 using genomic-based environmental surveillance.

## Introduction

In the past twenty years, several diseases caused by coronavirus have posed significant global health challenges-including Severe Acute Respiratory Syndrome (SARS, 2002), Middle East Respiratory Syndrome (MERS, 2012), as well as the current pandemic of COVID-19 [1,2]. COVID-19 is caused by a single-stranded, positive-sense RNA virus (SARS-CoV-2) which belongs to the *Coronaviridae* family [2]. The outbreak that was first detected in Wuhan (China) was declared a pandemic by the World Health Organization (WHO) on 11 March 2020, as it rapidly spread in over 114 countries [3]. As of January 14 (2021), over 108 million cases have been reported worldwide and have claimed over 2.3 million deaths [4]. In Nepal, there are 272,840 confirmed COVID-19 cases and 2055 deaths [5]. Gastrointestinal (GI) symptoms are often common in patients infected with SAR

S-CoV-2, with one hospital in the US reporting 70% of GI patients testing positive for coronavirus [6–9]. Although the primary source of transmission is through respiratory aerosol, studies have confirmed fecal shedding [10,11] and potential fecal-oral transmission of coronavirus [12,13]. Since SARS-CoV-2 is shed through feces and have also been detected in wastewater [14,15], detecting the virus in sewage and wastewater can serve as an early detection method for identifying communities with circulating virus in densely populated cities.

Environmental surveillance (ES) offers a complementary and more feasible approach to clinical disease surveillance. This approach provides unbiased information on a population level and detects viral shedding by symptomatic and asymptomatic patients [1,16], thus providing a snapshot of the outbreak over an entire sewage catchment area and an early indication of clinical cases in the area [17]. Because not all symptomatic patients get tested due to reluctance or lack of access to tests, clinical samples are not a sensitive indicator of the occurrence of cases in a given area [18,19]. As experienced by many countries, clinical and community-based COVID-19 surveillance is expensive, technically challenging, and time-consuming, and therefore hard to implement in the manner needed to rapidly inform public health measures to contain the outbreak [10,20,21]. A longitudinal study conducted in Boston (USA) between March-April in 2020 showed a high correlation between environmental samples testing positive for the virus 4-10 days before symptoms presented in people in the sampled areas [16]. Wastewater sampling was as sensitive as stool sampling for viral detection during vaccine-derived poliovirus outbreaks in Cuba [22]. The versatility of environmental samples combined with the implementation of cheaper, user-friendly, and short turnaround sequencing tools such as MinION (Oxford Nanopore Technologies, UK) [23] will be invaluable in rapidly identifying viral strains and detecting clusters of infection going forward.

Whole-genome sequencing (WGS) of viral pathogens is frequently used in disease surveillance and monitoring, and the importance of genomic surveillance has been widely recognized as an important method of understanding viral evolution [24]. Portable and reliable sequencing tools such as MinION can play a crucial role in conducting genomic surveillance in a developing country like Nepal where access to high throughput DNA sequencing machines is limited. The pocket-sized and field-deployable, the next-generation sequencer has enabled real-time outbreak surveillance of several recent outbreaks of Zika, Ebola, and Lassa viruses [1]. Obtaining whole or partial genome sequences of the virus during outbreak is critical in understanding viral evolution and disease epidemiology, and can help design accurate diagnostic tests for rapidly evolving RNA viruses such as SARS-CoV-2 [25].

This study was conducted in some selected areas of Kathmandu (Nepal) to evaluate the effectiveness of SARS-CoV-2 detection and characterization from environmental samples using portable DNA sequencing technology (MinION). Studies in other countries have detected SARS-CoV-2 in environment samples collected from wastewater treatment plants (WWTP) [2,16,26– 30]. However, Kathmandu has only one functional WWTP (out of five) and therefore, collecting all representative citywide sampling from WWTP is not possible. Since we had mapped sewage lines in some parts of the city for our ongoing surveillance of typhoid using environmental samples, we used the same sampling sites to conduct SARS-CoV-2 environmental surveillance. This cross-sectional study successfully detected, quantitated, and characterized the SARS-CoV-2 virus from environment samples collected from the wastewater outlets and utility holes within catchment areas, providing important information on the circulating strains of the virus in some communities of Kathmandu.

## Materials and Methods

### Sewage sample collection and processing

Much of the field sampling strategy was derived from our ongoing environmental surveillance work to determine the incidence of *Salmonella typhi* (the pathogen that causes Typhoid fever) in the Kathmandu valley, with an estimated population of over 4 million people [31]. As part of that study, we had comprehensively mapped sewage lines in the Lalitpur area of the Kathmandu valley. Also, we conducted field surveys to map out population size based on catchment areas. For our SARS-CoV-2 environmental surveillance, we used these sites and identified the catchment areas (n=20).

The initial feasibility and optimization study (May 13-July 5, 2020) was carried out with sewage samples collected around a hospital (Armed Police Force Hospital-APFH) with known COVID-19 patients. Three different sites (Shankhamul (TH-1, N27.678734/E085.334175), Thapathali (TH-2, N27.686754/E085.325203), and Teku (Te-3, N27.693163/E085.305504)) in the Kathmandu valley were also selected based upon catchment area coverage and representative population density.

Using an optimized protocol for detection and characterization of SARS-CoV-2, sewage samples were collected for the environmental and genomic surveillance (July 26-December 1, 2020) of three wards (Ward 9-Balkumari; Ward 11-Sankhamul/Jwagal, and Ward 17-Gwarko) located in the Lalitpur Metropolitan city of the Kathmandu valley (Nepal).

Sewage samples were collected using an automated robotic pump (Biobot Analytics Inc., Cambridge, USA). For protocol optimization, a total of 500ml of a composite (20.8 ml/hr) sample of 24 hours [16] was collected from the control site (APFH). But for the environmental surveillance, grab (50 ml) sample was preferred based upon outcome of optimization [34]. The probable reason for the consistent result in grab samples compared to the composite samples could be the dilution of viral particles by sewage discharge of non-viral peak hour [34]. The grab sewage samples were collected during the peak hour from 7:00AM to 9:00AM [35] based on the assumption that toilet activity and sewage discharge would be high during that time period.

The samples were transported immediately to our Kathmandu-based laboratory using cold chain for processing. Each sample was pasteurized at 60°C for 90 minutes in a water-bath [16] to inactivate any virus. Pasteurized samples were then subjected to differential centrifugation for virus segregation from sewage sludge by centrifuging at 3000 rpm for 30 minutes to pellet the bacterial cells and debris. The pellet was discarded, and the supernatant was precipitated for virus recovery using 15% PEG-6000 and 2% NaCl and gently shaken for 24 to 48 hours at 4°C [36]. The viral precipitates were then pelleted at 8000 rpm for 40 minutes and the pellet was re-suspended in 400µl of SM buffer (Tris/NaCl/MgSO4) for further processing (Fig 2). The samples were handled in enhanced Biosafety Level 2 laboratory with full personal protective equipment.

**Fig 1.**
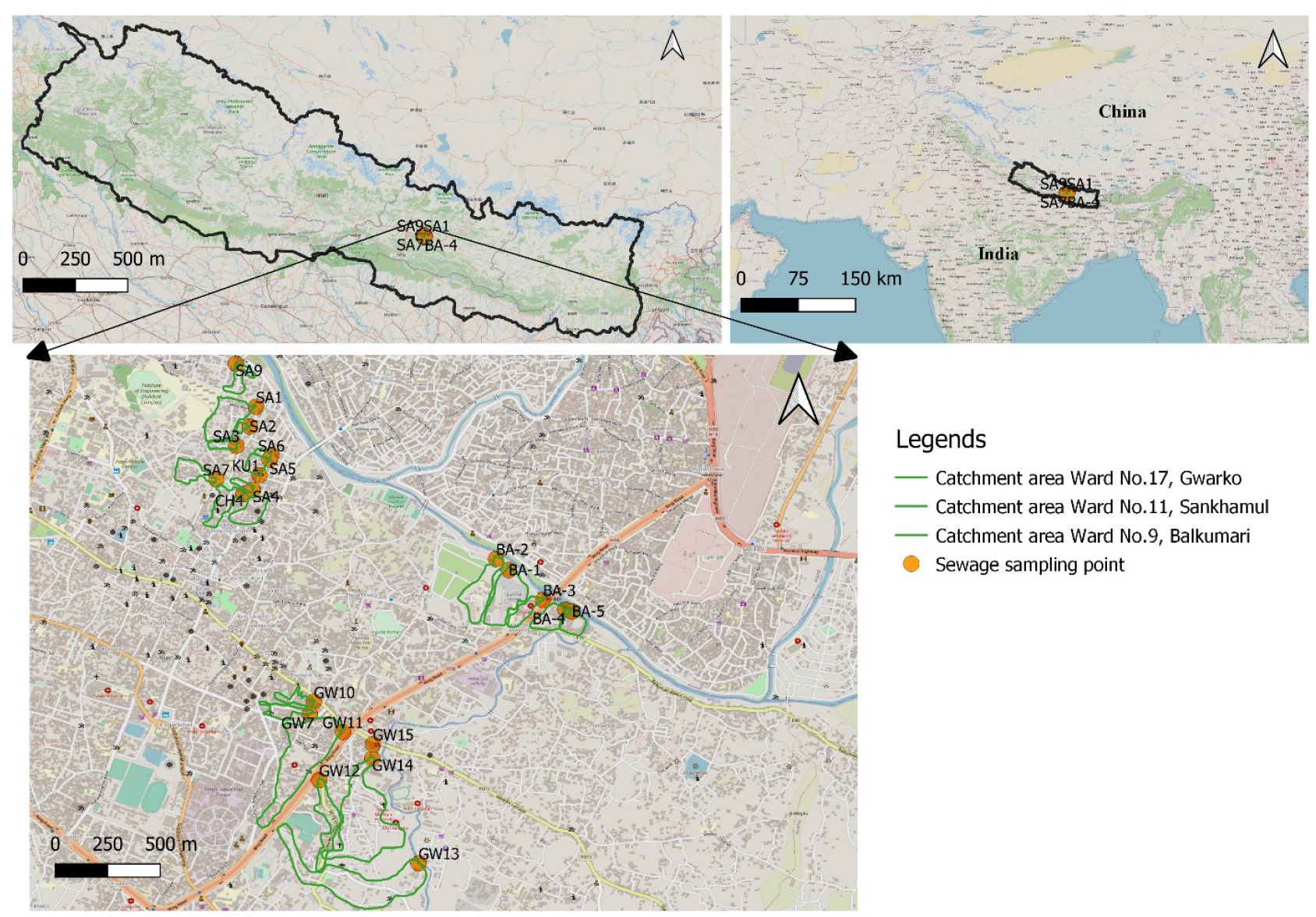
Map of the sampling sites and respective catchment areas selected for the SARS-CoV-2 environmental surveillance in the Kathmandu valley. The sites were visualized in a map using QGIS v3.16 [32] against a base map obtained from the Open Street Map [33].

**Fig 2.**
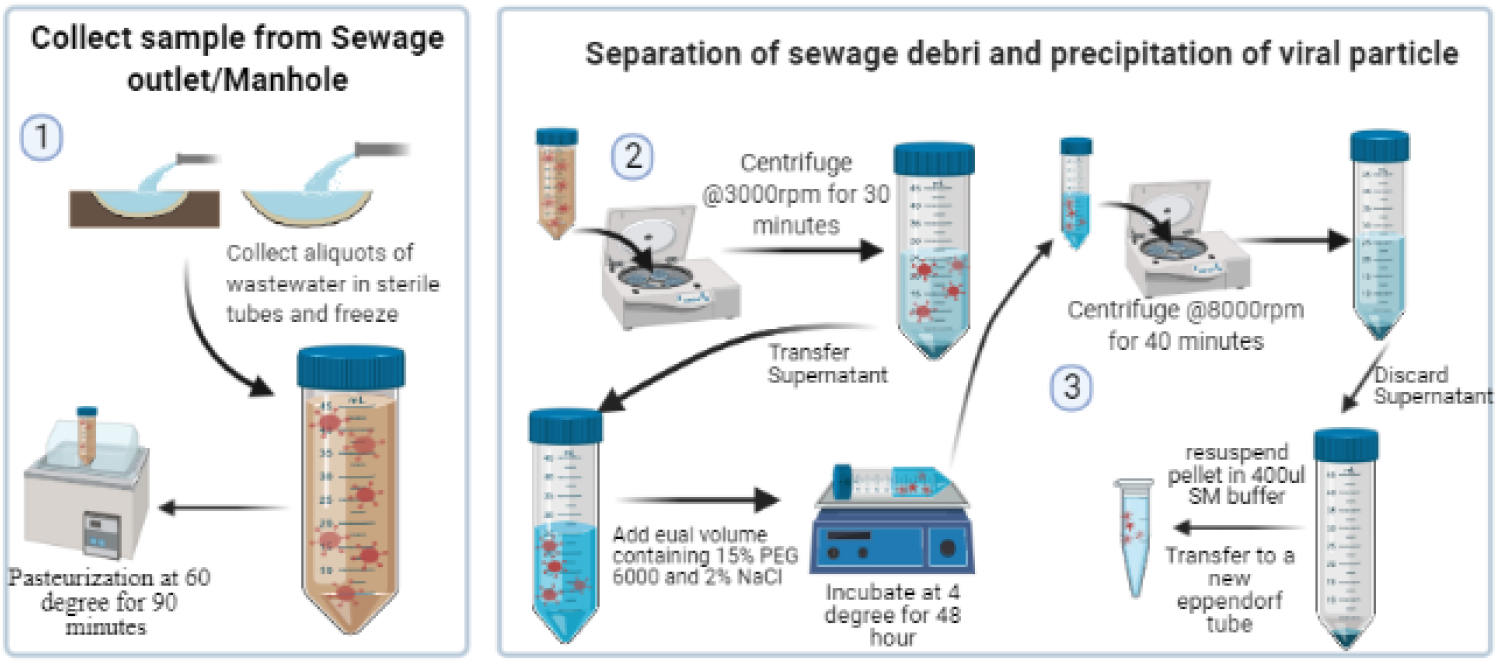
Environmental (sewage) sample processing for viral RNA extraction-. 1) grab sewage samples collected, 2) differential centrifugation for virus isolation, 3) supernatant precipitation and virus recovery. This illustration was created using BioRender app [37].

### SARS-CoV-2 detection using RT-PCR assay-Primary screening

We took samples from the sewage pipes of a selected hospital (Nepal Armed Police Hospital) with known admitted COVID-19 patients as a positive control site. Initially, regular diagnostic Real-Time polymerase chain reaction (PCR)-based assay (detecting envelope, nucleoprotein, and ORF1) was used to detect and quantify (sensitivity ∼20 viral copies/ml) SARS-CoV-2.

We also used nested PCR-based assay (which detects all coronavirus, Fig 3) followed by meta-barcoding to increase the sensitivity significantly and confirm the presence of other circulating coronaviruses along with SARS-CoV-2.

**Fig 3.**
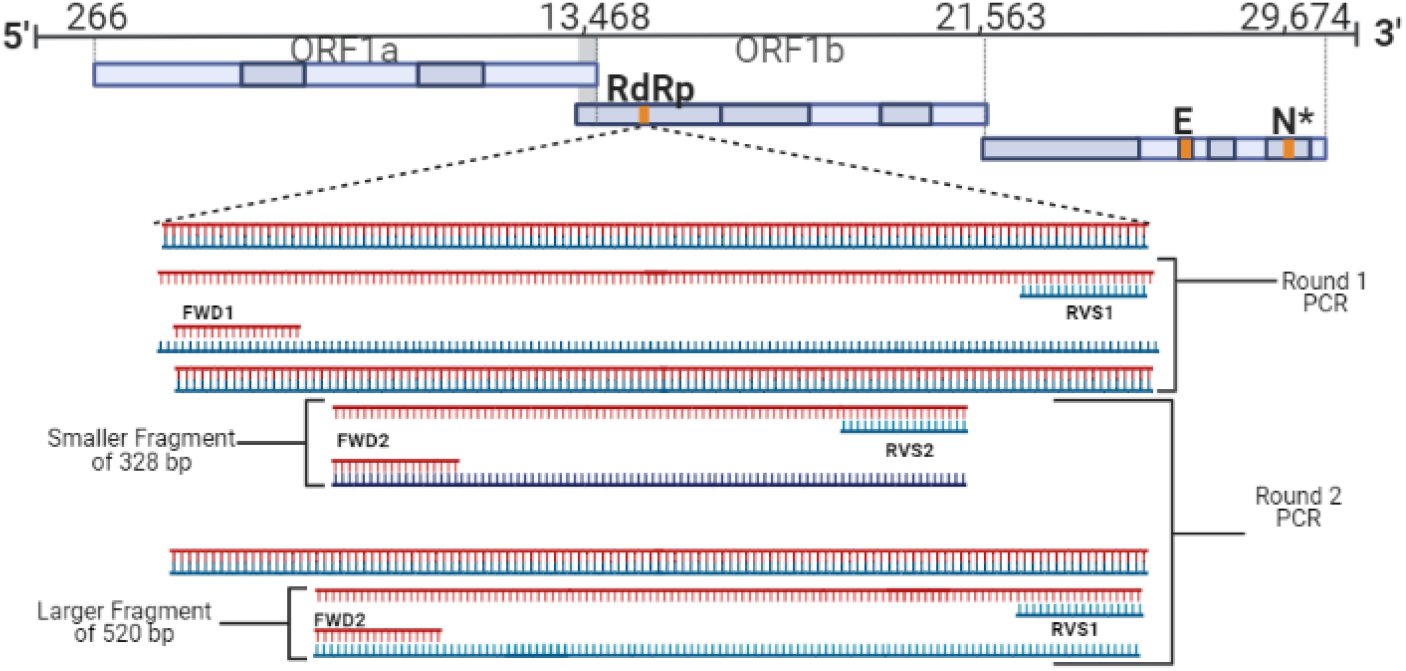
**Nested PCR to detect corona virus-using two sets of primers (FWD1/RVS2 & FWD2/RVS1) two PCR amplicons (328bp and 520bp) were produced which were cleaned and sequenced using Flongle MinION**. The obtained sequences were then BLAST analyzed in the NCBI database for viral taxonomic identification [38]. The images were created using BioRender app [37].

RNA was extracted from the viral suspension (200µl) (abGenix viral DNA and RNA Extraction Kit, AIT Biotech, Singapore) in an automated DNA and RNA extraction system (abGenixAIT Biotech, Singapore). SARS-CoV-2 was detected in sewage samples using Allplex™ RT-PCR SARS-CoV-2 Assay (Seegene Inc., Korea). The 5µl of extracted RNA was mixed with 10µl of qPCR mix consisting 2019-nCoV MOM (3µl), Real-time One-step Enzyme (1.2µl), 5X Real-time One-step Buffer (3µl) and Nuclease free water (2.8µl) with manufacture recommended conditions-cDNA synthesis (50°C for 20 min); polymerase activation (95 °C for 15 min); PCR (45 cycles of denaturation at 94°C for 15 s, and annealing/extension at 58°C for 30 s). 2019-nCoV_RdRp (ORF1ab) Positive Control (cat. 10006897, IDT USA) obtained from the University of California-Davis with known copy number was used to build a standard curve to determine the viral load of SARS-CoV-2. The viral load quantitation data of each site were visualized geospatially using the Heat-map plugin in QGIS v3.16 [32]. The viral load data were first converted to log scale, and then attributes were used to generate the heat-map, which was plotted against a base map obtained from open street maps.

### Coronavirus detection using meta-barcoding in MinION

To detect the presence of coronavirus other than SARS-CoV-2, we further screened RT-PCR negative samples with corona viral family-specific nested PCR modified on Quan et al. protocol using primers sets (FWD1/RVS2 &FWD2/RVS1) and producing 328bp and 520bp PCR amplicons [39] (Fig 3). 8µl of RNA template was mixed with 1µl of random hexamers and 1 µl of dNTPs and incubated at 65°C for 5 minutes. This mixture was combined with 10µl of synthesis mix consisting 10X RT (2µl), 25mM MgCl2 (4 µl), 0.1M DTT (2µl), RNase Out (1µl) and SSIII (1µl) (Superscript III (SSIII) cDNA synthesis kit, Invitrogen, USA). The final mixture was incubated at room temperature for 10 minutes, followed by incubation at 50°C for 50 minutes, and terminated at 85°C for 5 minutes. To remove any remnant RNA, the mixture was treated with 1µl of RNase H and incubated at 37°C for 20 minutes. The PCR amplicons were purified using a Montage Gel Purification kit (EMD Millipore Corp, USA) and sequenced using a portable next-generation sequencing device (Flongle MinION, Oxford Nanopore Technologies, UK). Nanoplot v1.30.1 [40] was used to check the quality of the raw nanopore fastq reads, and adapter sequences were trimmed using Porechop v0.2.4 [41]. Length and quality filtering were performed using Filtlong v0.2.0 [42], with minimum length and q-score thresholds of 300bp and seven. The cleaned sequence reads were then de-novo assembled using Canu v2.0 [43] to generate contigs with an overlap parameter threshold of 100bp. The scaffolds generated were subjected to the Basic Local Alignment Search Tool (BLAST) [44] for taxonomic identification against locally downloaded GenBank database (Release 240, October 15 2020).

### Whole-genome sequencing of SARS-CoV-2 using MinION

#### Library preparation and sequencing using MinION

Tiled amplicon sequencing was used to amplify the whole genome of SARS-CoV-2 using ARTIC PCR protocol [45]. This protocol has been widely adopted to sequence the entire genome of SARS-CoV-2 from clinical samples. With a set of 98 PCR primers, ARTIC PCR amplifies ∼400bp amplicons spanning the SARS-CoV-2 genome with adjacent ∼200bp overlaps. The sequencing library preparation is done by pooling these 98 primers in two pools and running two PCR reactions. We expect environmental samples to be highly degraded, and hence this protocol suits well for our study as smaller-sized amplicon increases the success of whole-genome amplification.

25µl reaction volume from each PCR pool were prepared containing 12.5µl Ampligold 360 2X MM, 3.6µl of primers and 2µl template cDNA. The PCR conditions were: 95°C for 30 secs followed by 45 cycles of 95°C for 15 secs, 63°C for 5 mins. The ARTIC PCR amplicons were further cleaned (AMPure bead, Agencourt, Beckman Coulter, USA) and quantified (Qubit 3, Invitrogen Thermofisher Scientific, USA). The PCR amplicons were pooled into a single tube at an equimolar concentration of 100fm. Native barcodes were assigned for each sample using Native barcoding Expansion kit (EXP-NBD104) and ligation sequencing kit (SQK-LSK109) and run in Flongle Flowcell using MinION (Oxford Nanopore, UK). Up to 14 samples were multiplexed in three FLO-FLG001 Flongle flow cells (R9.4.1) and sequenced on MinION Mk1B device in three different sequencing runs.

### Bioinformatics workflow for consensus genome sequence generation

MinKNOW software (Oxford Nanopore, UK) was used to monitor sequencing run, collect the raw data, and perform real-time base calling. The RAMPART v1.2.0 [46] software package developed by the ARTIC network was used to visualize read mapping and genome coverage in real-time for each barcode.

Data analysis was done using the ARTIC pipeline v1.1.3 [47] of the ARTIC network’s nanopore bioinformatics protocol [48]. Base-calling was performed in real-time by Guppy v4.4.0 (Oxford Nanopore, UK) integrated within MinKNOW v20.10.3 (Oxford Nanopore, UK) using high-accuracy base calling mode in Thinkpad P72 Mobile Workstation (Lenovo, USA), which generated fast5 and fastq reads.

De-multiplexing was performed by Guppy using strict parameters requiring barcodes (indexes) at both ends of the reads and quality filtering (qscore threshold 7) was performed within MinKNOW platform. This generated pass and fail reads for each barcode in individual directories.

Further filtering of read length was performed using the ARTIC pipeline with *guppyplex* option considering parameters of certain threshold length only (range 400 bp −700 bp) that removed obvious chimeric reads.

The processed reads were subjected to mapping, aligning, variant and consensus calling using the ARTIC pipeline with the *minion* option. The reads were mapped against the SARS-CoV-2 reference (NC_045512/MN908947) using the Minimap2 v2.17 [49]. The aligned read files were sorted using SAMtools v1.11 [50] to obtain coverage data and a consensus sequence.

### Whole-genome sequencing of SARS-CoV-2 using MiSeq

Using identical ARTIC amplicons, the first two positive samples from the study were sequenced in MiSeq (Illumina, USA) platform. The amplicons were cleaned (AMPure bead, Agencourt, Beckman Coulter, USA), and a library was prepared using the Nextera XT library preparation kit [24]. The data was processed and a consensus sequence was generated using a nextflow-based SARS-CoV-2 Consensus Genome Pipeline developed by Chan Zuckerberg Biohub [51].

### Geo-spatial mapping of identified variants of SARS-CoV-2

Identification of known and novel mutations in the obtained whole-genome sequences of SARS-CoV-2 was performed using a CoV-GLUE web application-based bioinformatics tool [52]. This tool is based on a GLUE data-centric bioinformatics environment that analyzes nucleotide variations in user-submitted sequences of SARS-CoV-2, enabled by data from EpiCoV of GISAID database. The tool compares and analyses the sequences against the reference sequence of SARS-CoV-2 and generates lists of amino acid replacements and coding region indels. The mutation table data obtained from CoVGlue was formatted to contain column names as follows; “sample”, “gene”, “variant_class”, “amino.acid.change”. Once the data has been formatted, it was imported to R studio and were processed using plugin GenVisR to generate mutation landscape plot (Waterfall plot) [53].

The tool also provides a lineage based on Pangolin classification of SARS-CoV-2 clades and its LWR (likelihood weight ratio) score, which approximates confidence level of clade assignments based on quality of the submitted sequence. The distribution of lineages across the sampling sites were visualized in a map using QGIS v3.16 [32] against a base-map obtained from the Open Street map [33].

## Results

### Optimization of protocol for the detection of SARS-CoV-2 and other coronaviruses in sewage samples

Four preliminary sites were selected (Armed Police Force Hospital (APFH), Thapathali (TH1), Shankhamul (TH2) and Teku (Te3)) for the initial protocol development and optimization. Along with SARS-CoV-2, we also detected other coronaviruses, including Human coronavirus 229E, Duck-dominant coronavirus, Dromedary camel coronavirus HKU223, and Rat coronavirus, in two preliminary sites (TH1 and Te3) (S1 Table).

### Whole-genome sequencing of SARS-CoV-2 using MinION and MiSeq

Whole-genome sequencing data of detected SARS-CoV-2 were obtained using MinION and MiSeq from two different sites (TH1 and Te3). The result showed a slight difference in genome coverage and contigs recovered from both the platforms. We observed slightly higher genome fraction coverage on MiSeq than on MinION. In one of the samples (TH1), MiSeq produced relatively high (47.2%) genome fragment sequences compared to MinION (32.2%). A similar result was observed in the other sample (Te3), with MiSeq producing around 42.9% sequences compared to 31.1% from MinION (Table 1).

**Table 1.** MinION sequencing run details and run statistics.

|  | **First Run** | **Second Run** | **Third Run** |
| --- | --- | --- | --- |
| Number of samples | 2 | 6 | 6 |
| Total raw reads | 693,554 | 312,602 | 128,107 |
| Total mapped reads | 414,002 | 253,168 | 102,011 |
| Median reads length | 482 | 494 | 477 |
| Passed reads | 433,024 | 258,256 | 104,856 |
| Genome fraction (%) at >20x | 33 to 36 | 32.3 to 51.9 | 28.2 to 71.8 |
| Run duration hours | 23.55 | 18.47 | 14.89 |

CoVGlue analyses [52] of consensus genomic sequences from both the sequencing platforms indicated the presence of the same Pangolin lineage (**B.1**). Major mutations were plotted to visualize and differentiate key differences between MiSeq and MinION using the GenVisR tool in R studio Version 1.4.1103. We detected **D614G** which is the defining mutation of the G clade [54]. MiSeq did detect more mutations than MinION (Fig 4). This finding can be due to greater sequencing depth (1 million reads) obtained in MiSeq than in MinION using Flongle (378,000 reads). Despite shallow sequencing depth in MinION, both platforms successfully detected major mutations and provided adequate sequencing coverage data for lineage identification [55].

**Fig 4.**
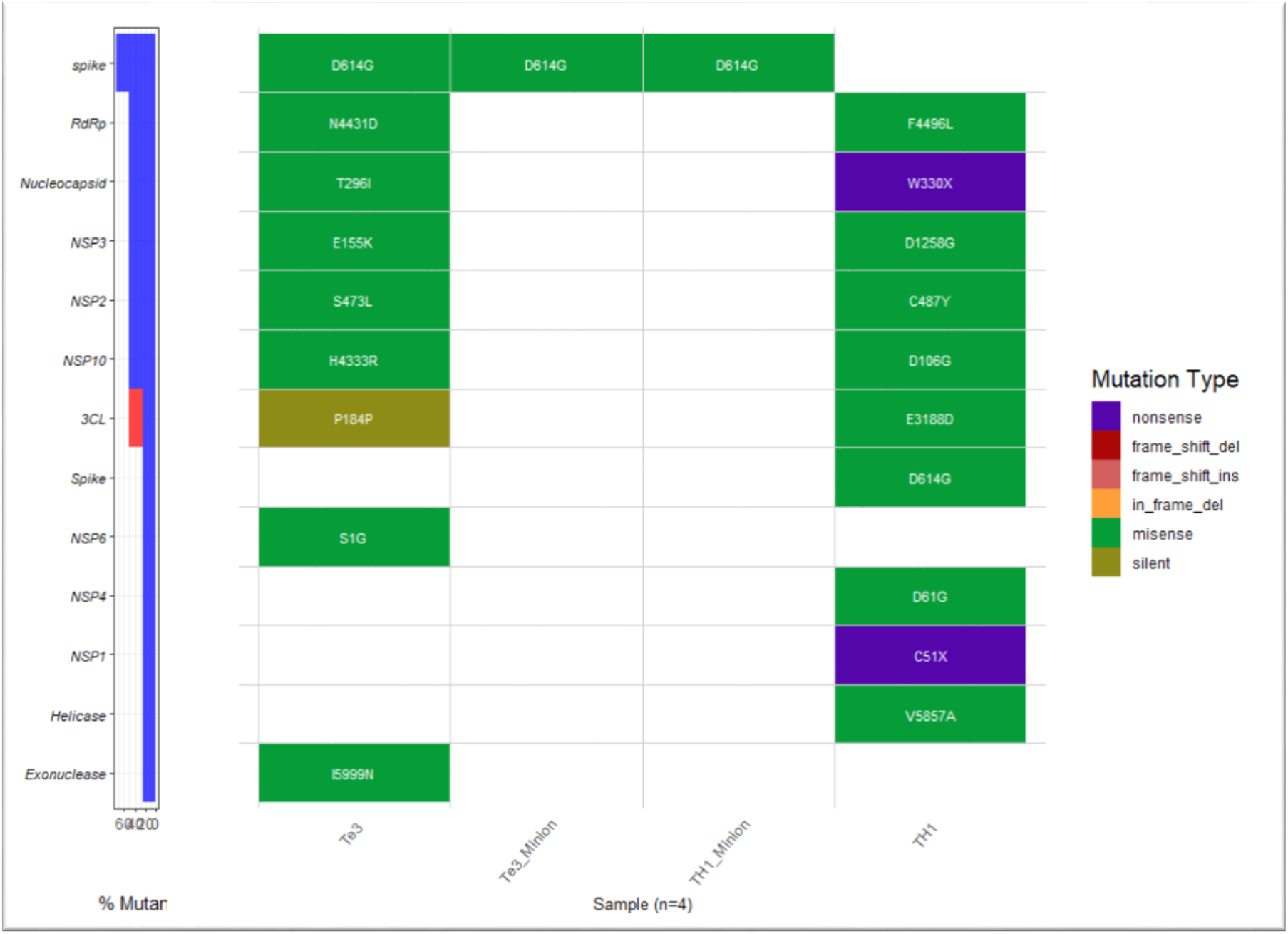
Detected mutations in the environmental samples using MinION and MiSeq. In this waterfall plot, rows represent genes with key detected mutations with the columns’ corresponding samples. Based on the weight of mutation type (orderly displayed in the legend with **nonsense** having greater impact and **silent** having the least impact on gene expression), only major mutations in a particular gene in each sample are displayed. Samples from two sites (TH1 and Te3) were sequenced using MiSeq (TH1 and Te3), and MinION (TH1_Minion and Te3_Minion) (Waterfall plot created using CovGlue and GenVisR [56] on R studio version 1.4.110).

### SARS-CoV-2 genomic-based environmental surveillance with sewage samples

Out of 20 sites, sewage samples from the 16 sites had detectable SARS-CoV-2 (S2 Table). A heat-map of all these 16 SARS-CoV-2 positive sites was created based on viral load intensity and corresponding geospatial data. With a relatively large number of sampling points, dense populations and detected viral load (295 copies/ml), Ward 11 (est. pop=4632) is colored dark in the heat-map. Although Ward 17 (est. pop=9500 people) had the most intense viral load (242,000 copies/ml), the sampling points do not cover the entire area, and hence lightly colored in the heat-map than Ward 11. In Ward 9 (est. pop= 2791 people), we did not detect SARS-CoV-2 (Fig 5).

**Fig 5.**
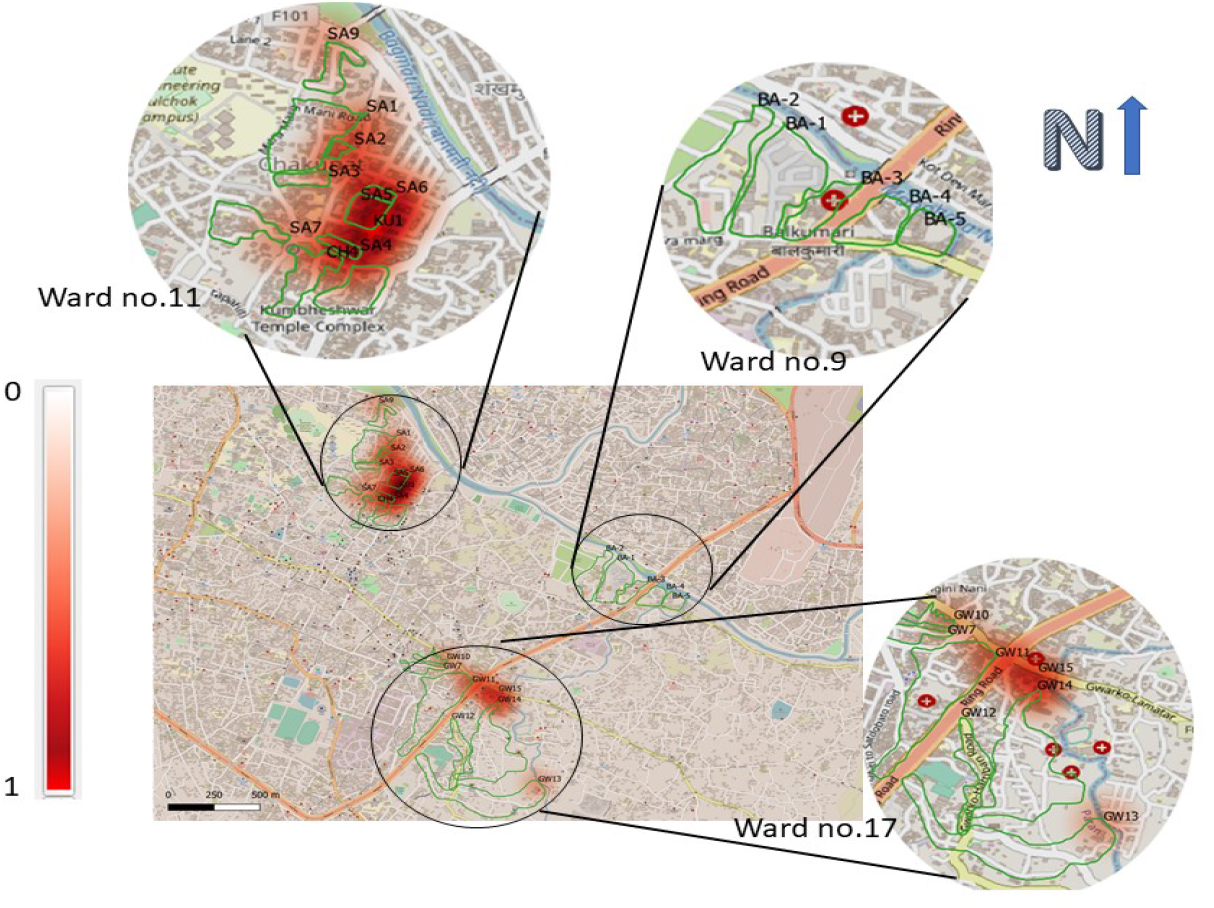
SARS-CoV-2 heat-map based on viral load intensity from sites with detectable virus. Heat-map of three sampled wards (11, 9 and 17) of the Lalitpur Municipality (Kathmandu valley) with SARS-CoV-2 presence and its viral load intensity in sewage samples. The map was created using QGIS v3.16 [32] with base-map from the Open Street Map [33].

### Whole-genome sequencing of SARS-CoV-2 on MinION

Only samples from 14 sites (GW11, SA2, SA7, CH4, KU1, SA5, GW10, GW13, GW14, SA6, SA4, SA9, GW12 and GW15) yielded PCR amplicons using ARTIC primers, of which only 12 samples (GW11, SA2, SA7, CH4, KU1, SA5, GW14, SA6, SA4, SA9, GW12 and GW15) had enough quantifiable PCR products (S3 Table, S4 Table) and were tagged with Native Barcode from NBD01 to NBD012 [24]. The consensus genomic sequences of SARS-CoV-2 have been submitted to National Center for Biotechnology Information (NCBI) GenBank (accession no. MW739929-MW739930). MinION sequencing was performed with 6 samples at a time, and run data is shown in Table 1.

All of the consensus sequences obtained from MinION were compared against the reference sequence (accession no. MN908947.3) using the CovGlue web application. **B.1** (16.7%), **B.1.36** (33.3%) and **B.1.1** (50.0%) were identified as circulating lineages of SARS-CoV-2 in the environmental (sewage) samples (Fig 6).

**Fig 6.**
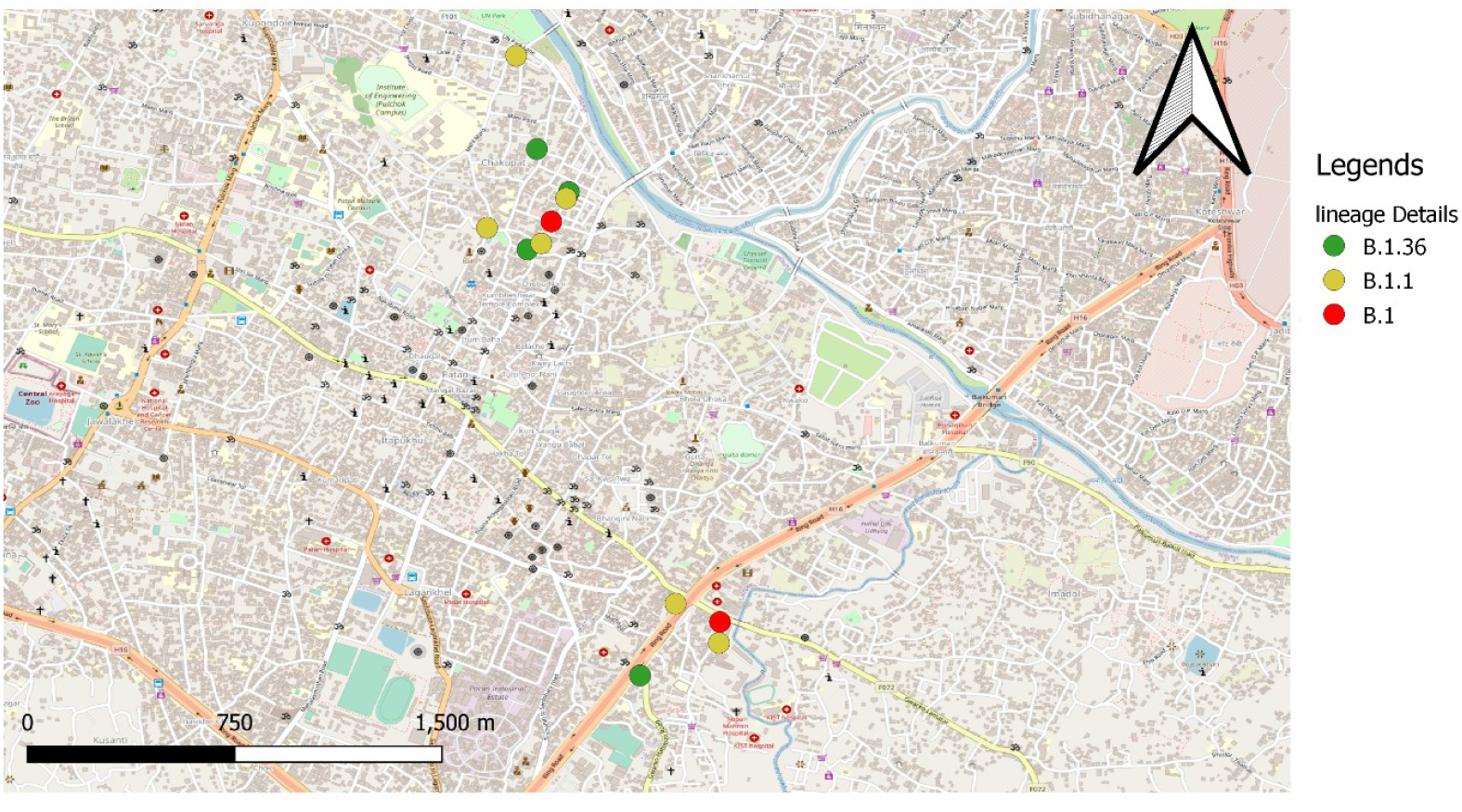
Geographic distribution of the detected SARS-CoV-2 Pangolin lineages *[56]* in the environmental samples from CovGlue submission. Three wards of Lalitpur Municipality (Kathmandu valley) with a distribution of detected SARS-CoV-2 and its lineages in sewage samples (Created using QGis 3.16 using Open Street Map [33]).

### SARS-CoV-2 evolution trend

Transition-Transversion mutation summary was visualized with a waterfall plot and created with GenVisR [55] (Fig 7). Nucleotide mutation summary of all the samples showed a greater frequency of transition (G>A (G>U) or C>T) in most of the samples, followed by transversion (G>T or C>A (C>U). Overall transition seems to have occurred in greater frequency than transversion (transition=57.5%, transversion=42.4%); and A>C or T> G transversion is rarely detected (16.4%) (Fig 8). A higher number of transitions in detected mutations indicate a rapid evolution of SARS-CoV-2 in the host [57].

**Fig 7.**
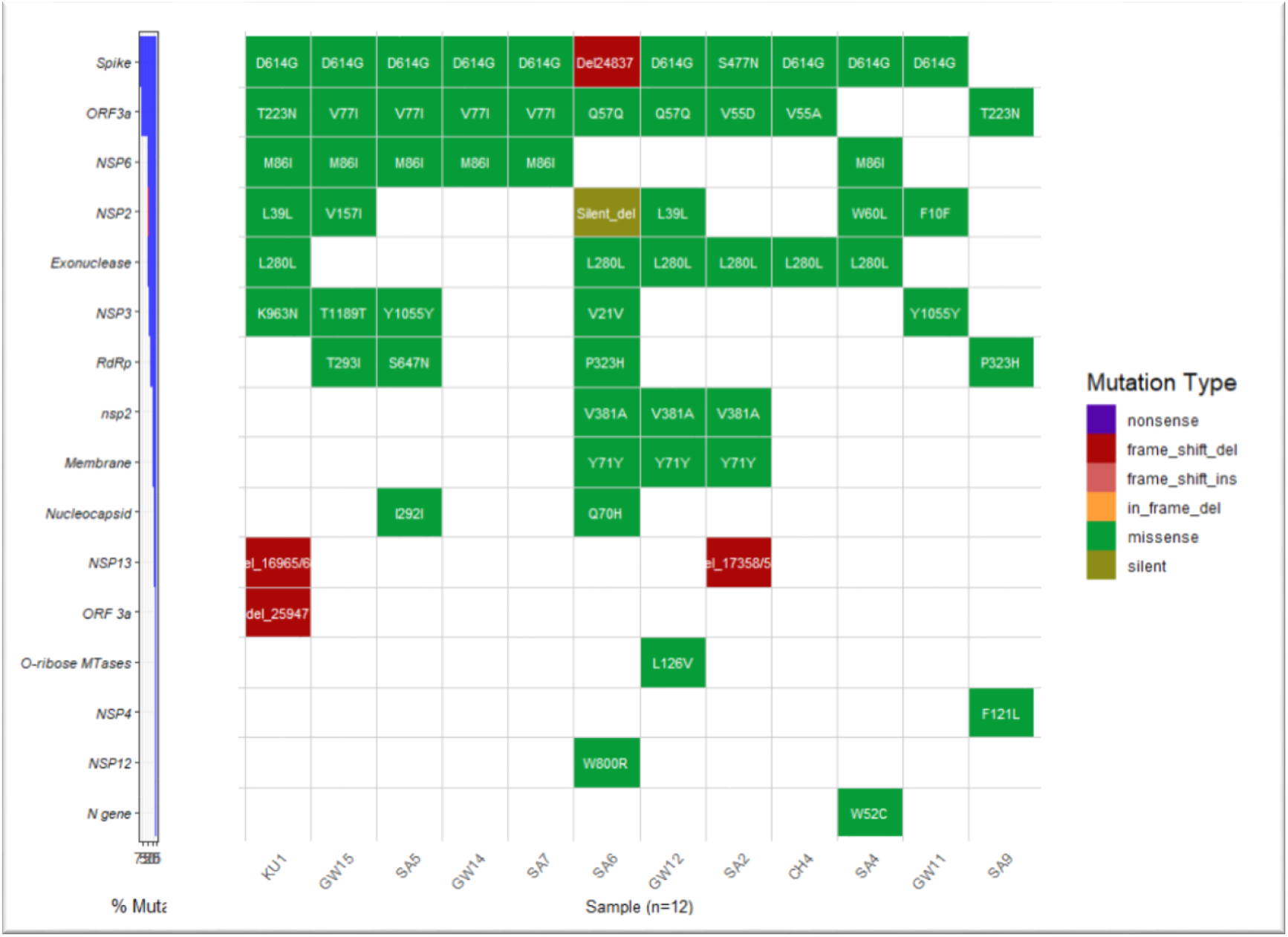
Detected SARS-CoV-2 genes and their associated mutations. Rows represents genes with major mutations detected while column represents sample, a waterfall plot was created based upon the weight of mutation type with nonsense having greater impact and silent having the least impact on gene expression. Created using CovGlue and GenVisR [56] on R studio version 1.4.110.

**Fig 8.**
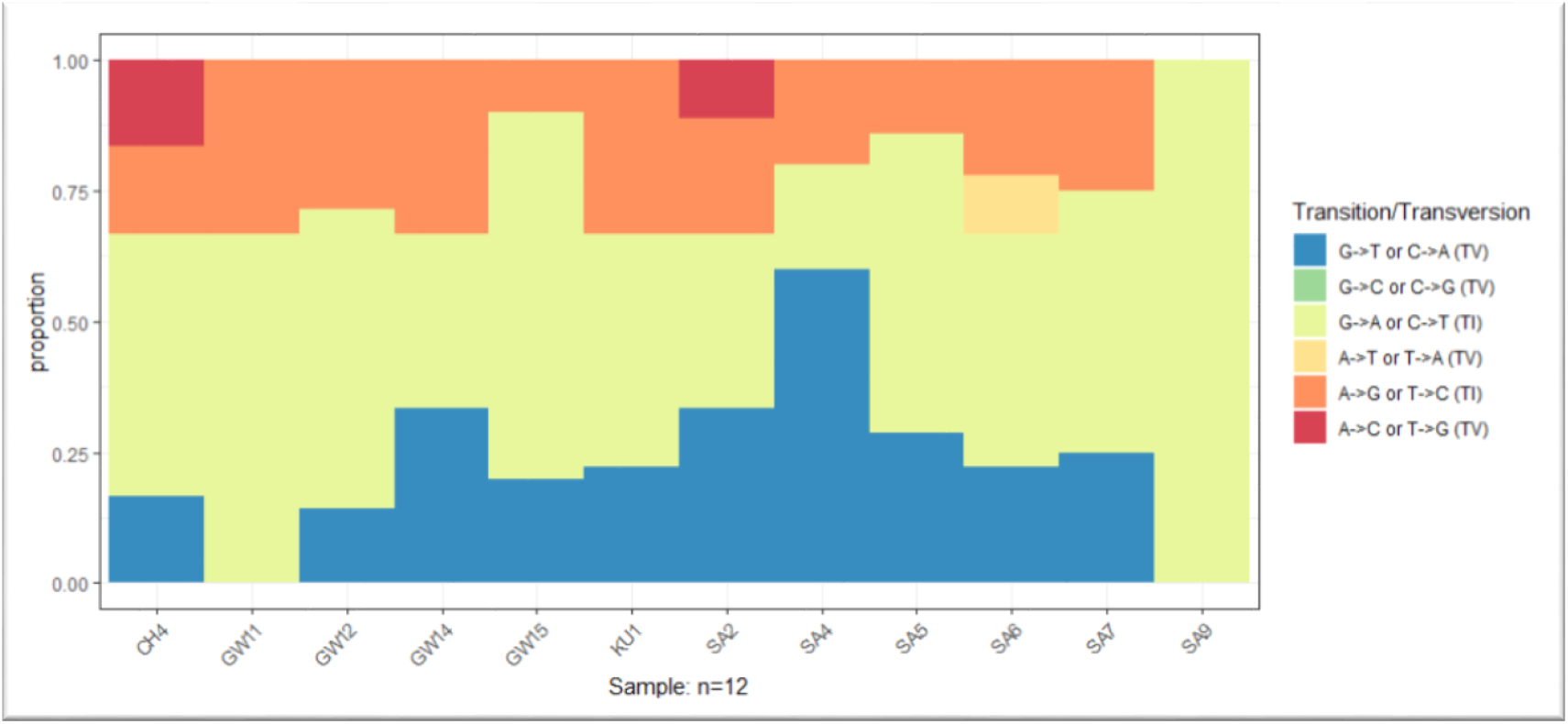
Transition-Transversion mutation summary plot from the mutation table detected by CovGlue using GenVisR [55] on R studio version 1.4.1103, showing various genes with detected mutations.

A total of 41 distinct mutations were detected across the SARS-CoV-2 genome with nine novel mutations - six due to deletion. Of all the deletions detected, two were non-codon aligning deletions, whereas the rest were frameshift deletion, including one in the spike gene (Fig 7, Fig 9). Major mutations were detected in spike, ORF3a, NSP6, and Exonuclease genes of SARS-CoV-2. **D614G** and **S477N** mutations were detected in the spike gene. Other genes with frameshift deletion include **NSP13** and **ORF3a** gene and one silent deletion on the **NSP2** gene (Fig 7, Fig 9).

**Fig 9.**
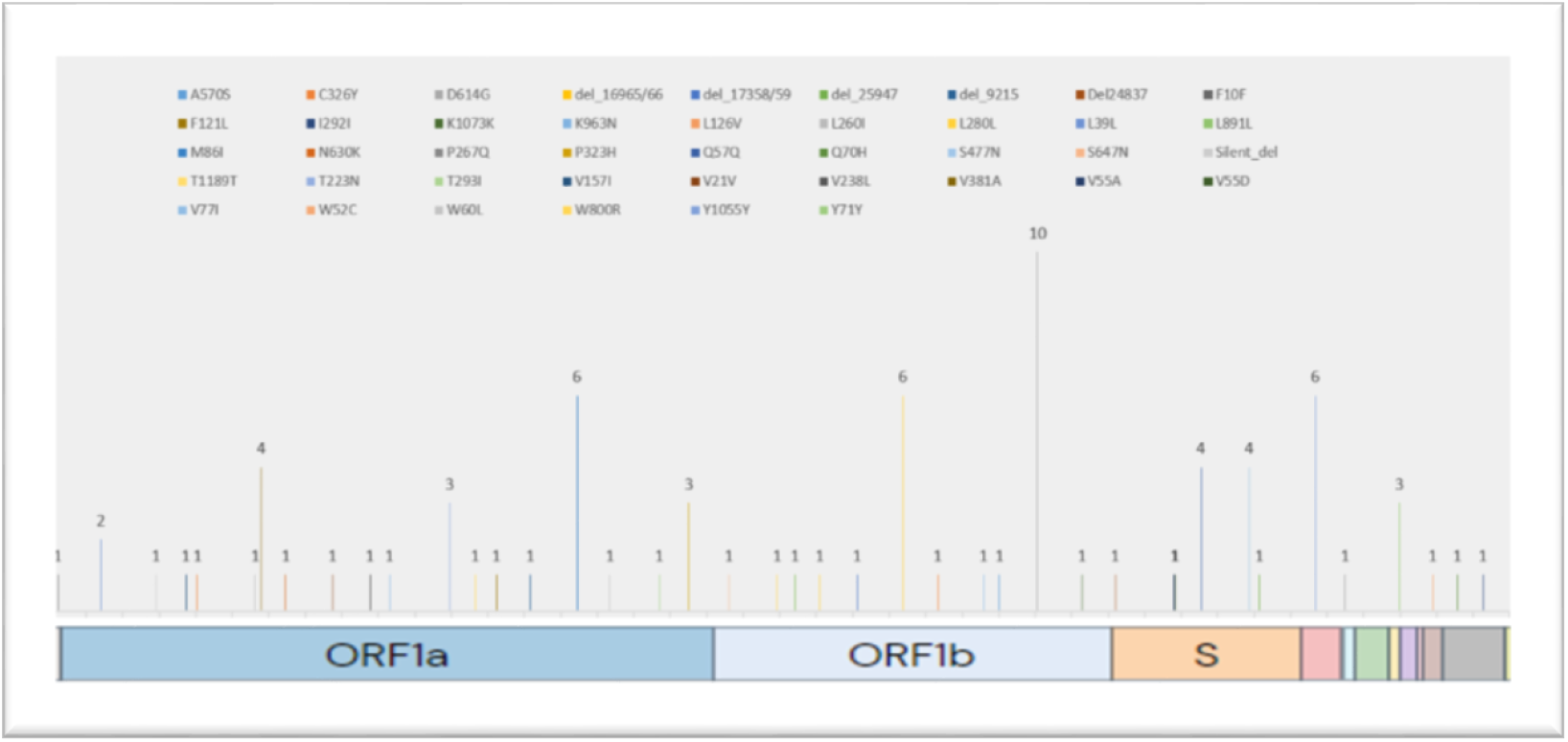
Detected mutations and their frequencies in the SARS-CoV-2 genome from environmental samples (n=12). The frequency of detected mutations on the corresponding genes is labeled at the top of the chart with height representing the frequency level. The lables on the X-axis were extracted from GISAID[58].

The novel mutations with amino acid changes were detected on the **W800R, W60L** and **W52C** in **NSP12, NSP2** and **N** genes. Among these three detected mutations, two were from the same sample (**SA4**), and one was from **SA6** collected from different sampling sites from the same ward.

## Discussion

As demonstrated by its successful application in the global efforts to eradicate poliovirus [24], ES can be an important supplement to clinical surveillance. In particular, ES can contribute to several critical areas, such as characterizing regional diversity in areas without clinical surveillance, mapping geographic micro-diversity to inform risk-based interventions, early detection of outbreaks before clinical presentation, and advancing real-time assessment of interventions to inform need for additional public health measures [24]. Compared to clinical surveillance, ES can be inexpensive and fast, but this approach has yet to be fully validated as a viable option for estimating disease burden in a field setting.

Rapid detection of SARS-CoV-2 in environmental samples (such as sewage) and viral strain characterization using portable genomic tools provide important information on disease distribution at a community level [24]. Such data can help identify transmission hotspots in urban and rural communities and provide information on circulating strains of viruses, an essential consideration for vaccination programs. We have optimized a system of detecting the virus from sewage samples, and based on viral load, we have constructed a heat-map of SARS-CoV-2 in the selected areas of the Kathmandu valley. In resource-limited, developing countries like Nepal, access to tools and technologies required to perform genomic-based detection and surveillance can be limited. This feasibility study has demonstrated that ES based on sewage samples using highly sensitive nested PCR combined with portable next-generation sequencing technology can provide adequate sensitivity, specificity, and sequencing depth to detect, quantify, and characterize SARS-CoV-2 accurately.

Whole-genome sequencing of the SARS-CoV-2 was obtained using ARTIC primers in two popularly used next-generation sequencing platforms (MiSeq (Illumina, USA) and MinION (Oxford Nanopore, UK)). One of our main objectives was to explore the feasibility of using a portable sequencing platform in Nepal. Using the same protocol, the sequencing data obtained from both these platforms were analyzed, and the results obtained were almost identical, demonstrating the suitability of using portable sequencing devices such as MinION to characterize viral (SARS-CoV-2) strains. However, with CoVGlue analysis, we were able to detect more variants (mutations) in Illumina MiSeq than ONT MinION using Flongle flowcell. We can use Flongle (MinION) for screening for rapid surveillance, and further detailed analysis can be done either on MiSeq or SpotON Flowcell (MinION).

In our ES samples, we were also able to detect other coronaviruses such as human coronavirus 229E, a known cause of the common cold with global distribution [24,59]. The presence of duck dominant coronavirus and rodent coronavirus in sewage suggests an abundant mixture of various coronaviruses originating from other species. A diversity of coronaviruses in sewage samples highlights the potential for emerging coronaviruses among species in close proximity in the urban setting, especially given the propensity for coronaviruses to recombine [60].

Out of 20 different sampled sites, 16 had SARS-CoV-2 present. The heat-map generated based on the intensity of viral load correlated well with the catchment area and population size (Fig 5). Analyzing enough sampling points can provide a good picture of circulating viruses in a community. Further studies can be designed to look into incidence estimation in communities and correlate that with available clinical data.

The obtained consensus genome sequences of SARS-CoV-2 from the sewage samples identified **B.1, B.1.36** and **B1.1** as circulating lineage. The lineage **B.1** was a strain circulating in the United States of America (46.0%), United Kingdom (13.0%), Canada (5.0%), Spain (4.0%), and France (4.0%); and was involved in the Italian outbreak [24,61]. **B.1.1** was also widely circulating around the globe (United States of America (27.0%), United Kingdom (22.0%), Canada (6.0%), Germany (6.0%), and Netherlands (5.0%)) and is cited as a European lineage [61]. Lineage **B.1.36** was the predominant strain circulating in the United States of America (94.0%), and is known as a USA lineage[61]. Whole-genome data of SARS-CoV-2 from 14 clinical samples have been sequenced at our facility for the Nepal Health Research Council (Government of Nepal). Data are publicly available in the GISAID database [24]. Among these, three samples originate from the Kathmandu valley, and identified lineages include **B.1.36** and **B.1.130**. The detected SARS-CoV-2 lineages (**B.1, B.1.1, B.1.36**) and key mutations such as **D614G** in the clinical samples were also circulating in the sewage samples.

Among the detected mutations, the majority were transitions that provide a snapshot of the evolutionary dynamics of SARS-CoV-2 in the host. Few novel mutations with possible clinical significance were also detected-such as frameshift and silent deletion mutations in **S, NSP13, ORF3a**, and **NSP2** genes. These novel mutations are yet to be reported in the global database, and their functional characterizations are further required. For example, frameshift deletion in the S gene could have important significance in clinical and epidemiological frames. The D614G (in S gene) was spontaneously detected in almost every sample, a defining mutation for the G clade [54]. This particular mutation is associated with increased infectivity of SARS-CoV-2 [23]. Another detected mutation, S477N (in S gene), has been reported to increase the binding affinity of the virus to the host’s angiotensin-converting enzyme 2 (ACE-2) receptor giving higher transmission capability [62,63]. Similarly, novel missense mutations detected in **NSP12, NSP2**, and **N** genes from sites in the same Ward indicated ongoing viral circulation in the community. These inferences tell us that the virus is spreading rapidly throughout the community, which provides the virus opportunities for rapid evolution. Such a scenario creates an environment for the emergence of new variants, which was also the reason behind the origins of emerging variants in the UK (B.1.1.7), South Africa (B.1.351), and South America (P.1) [64]. This finding also highlights the mandates for genomic surveillance to track viral evolution and emerging variants.

Three major vaccines (Pfizer-BioNTech, Moderna, and Oxford-AstraZeneca) approved by the WHO target spike protein to produce neutralizing antibodies against SARS-CoV-2 [65]. The detection of circulating viral variants through genomic surveillance has become more critical than ever to evaluate the efficacy of the vaccine against the evolving virus. For this reason, genomic surveillance is gradually being conducted in many developed countries, with the United Kingdom contributing actively to the global genomic database of SARS-CoV-2 [66]. However, developing countries are still struggling to understand COVID-19 incidences and transmission due to a lack of genomic laboratory capabilities. Genomic surveillance in environmental samples could be a possible solution for resource-strapped countries to identify and estimate SARS-CoV-2 presence at a community level and determine the circulating strains of the virus, which could be used to select the most appropriate vaccine.

## Data Availability

All data pertaining to this study is included in the manuscript.

## Acknowledgments

We would like to thank the Mayor’s Office of the Lalitpur Metropolitan City for providing us with permit and encouraging us to conduct this research. We would like to thank Professor Eric Alm and Dr. Noriko Endo of BIOBOT (USA) for providing composite sampling robots. We would also like to express our gratitude to the One Health Institute of the University of California-Davis and the USAID-funded PREDICT project for providing us with laboratory resources. Whole-genome sequencing was done at the Intrepid Nepal Genomic Center-some of our work was partially funded by the Australian Development Agency and PSI grant (the Netherlands). We would like to thank all these agencies for their support. And finally, we would like to thank everyone at Intrepid Nepal Pvt Ltd, Center for Molecular Dynamics Nepal and BIOVAC Nepal for their tireless work, even during nationally enforced lockdown in Nepal.

## Supporting Information Captions

**S1 Table:** Blast result analysis of Coronavirus origin from sewage samples of Pilot phase study.

| Sample sites | Query ID | Accession | Organism | Evalue | Seqlen | Querycov | Identity |
| --- | --- | --- | --- | --- | --- | --- | --- |
| TH1 | tig00000001 | AL158839.11 | Human | 1.00E-119 | 788 | 86 | 99.6 |
|  | tig00000002 | KT254285.1 | Duck-dominant coronavirus | 0 | 480 | 99 | 96.26 |
|  | tig00000003 | KF293666.1 | Human coronavirus 229E | 4.00E-78 | 466 | 44 | 92.96 |
|  | tig00000004 | MT655131.1 | SARS-CoV-2 | 0 | 472 | 92 | 98.17 |
|  | tig00000005 | KF294370.1 | Rat cov | 0 | 471 | 100 | 90.87 |
|  | tig00000006 | MN306040.1 | Human coronavirus NL63 | 0 | 391 | 100 | 99.49 |
|  | tig00000007 | no significant hits |  |  |  |  |  |
| Te3 | tig00000001 | LR824127.1 | SARS-CoV-2 | 0 | 422 | 96 | 95.82 |
|  | tig00000001 | KT254285.1 | Duck-dominant coronavirus | 0 | 466 | 100 | 96.15 |
|  | tig00000002 | KF294370.1 | Rat cov | 2.00E-123 | 316 | 99 | 92.04 |
|  | tig00000003 | no significant hits |  |  |  |  |  |
|  | tig00000004 | no significant hits |  |  |  |  |  |
| TH2 | tig00000001 | KT254285.1 | Duck-dominant coronavirus | 0 | 475 | 99 | 96.2 |
|  | tig00000002 | KF294370.1 | Rat cov | 1.00E-125 | 327 | 99 | 91.38 |
| APF | tig00000001 | MT628184.1 | SARS-CoV-2 | 3.00E-153 | 319 | 99 | 98.73 |

**S2 Table:** **Real time PCR data, viral load and population distribution of different sites. Amplified genes-envelope (E), internal control (house-keeping gene, IC), RdRp and nucleoplasmid (N) genes.**

| S.N. | Date | Sampling Site | RT-PCR Result (Ct value) |  |  |  | Viral Load (viral copies per ml) | Est. human pop. Community |
| --- | --- | --- | --- | --- | --- | --- | --- | --- |
|  |  |  | E gene | IC | RdRp | N gene |  |  |
| 1. | 26/7/2020 | BA-1 | - | - | 35 | - | - | 1486 |
| 2. | 26/7/2020 | BA-2 | 20.1 | - | - | - | - | 518 |
| 3. | 26/7/2020 | BA-3 | - | - | - | 36.4 | - | 636 |
| 4. | 10/09/2020 | GW13 | 33.9 | 33.5 | 36.1 | 35.5 | 2090 | Unknown |
| 5. | 27/09/2020 | GW10 | 32.9 | 29.9 | 37.3 | 34.7 | 1,950 | 532 |
| 6. | 4/10/2020 | GW7 | 36.2 | - | - | 34.5 | - | 330 |
| 7. | 9/11/2020 | GW11 | 25.9 | 27.6 | 29.5 | 27.5 | 242000 | 2367 |
| 8. | 11/11/2020 | GW14 | 28.0 |  | 30.1 | 28.4 | 43100 | Unknown |
| 9. | 12/11/2020 | SA1 | 34.0 | 35.6 | 36.2 | 33.8 | 295 | 286 |
| 10. | 18/11/2020 | SA2 | 29.0 | 30.5 | 30.5 | 29.6 | 28900 | 81 |
| 11. | 19/11/2020 | SA7 | 27.5 | 30.5 | 32.2 | 29.1 | 7840 | 756 |
| 12. | 20/11/2020 | CH4 | 30.6 | 30.3 | 33 | 31.0 | 3640 | 700 |
| 13. | 22/11/2020 | KU1 | 28.0 | 29.6 | 30.8 | 28.4 | 92,300 | 1686 |
| 14. | 23/11/2020 | SA6 | 32.3 | - | 34.7 | 32.0 | 321 | 101 |
| 15. | 24/11/2020 | SA5 | 27.8 | 30.2 | 28.8 | 28.1 | 60,400 | 400 |
| 16. | 25/11/2020 | SA4 | 28.4 | 30.6 | 34.2 | 29.8 | 2820 | 204 |
| 17. | 26/11/2020 | SA3 | 30.2 | 28.6 | 34.3 | 30.6 | 1640 | 191 |
| 18. | 27/11/2020 | SA9 | 30.4 | 28.6 | 33.7 | 31.1 | 2390 | 227 |
| 19. | 30/11/2020 | GW12 | 29 | 30.9 | - | 29.4 | - | Unknown |
| 20. | 1/12/2020 | GW15 | 32.7 | 26.3 | 33.1 | 32.6 | 3660 | Unknown |

**S3 Table:** **AMPure bead clean up and DNA quantification data of pool1 & pool2 artic PCR products in positive sewage samples before Indexing (Pilot study)**

| S.N. | Sample ID | ng of DNA present | Size of PCR product | Femtomoles of DNA present | Femtomoles of DNA required | ng of DNA required | Volume of DNA required | Volume of water required |
| --- | --- | --- | --- | --- | --- | --- | --- | --- |
| 1 | PL 1 Env5 | 4.86 | 400 | 18.72 | 200 | 51.92 | 10.7 | 39.3 |
| 2 | PL 1 Env10 | 0.85 | 400 | 3.27 | 200 | 51.92 | 60.7 | -10.7 |
| 3 | PL 1 E2 | 9.93 | 400 | 38.25 | 200 | 51.92 | 5.2 | 44.8 |
| 4 | PL 2 Env5 | 2.45 | 400 | 9.44 | 200 | 51.92 | 21.2 | 28.8 |
| 5 | PL 2 Env10 | 0.33 | 400 | 1.27 | 200 | 51.92 | 156.7 | -106.7 |
| 6 | PL 2 E2 | 0.51 | 400 | 1.96 | 200 | 51.92 | 102.7 | -52.7 |
| 7 | PL 1 env1 | 33.60 | 400 | 129.43 | 200 | 51.92 | 1.5 | 48.5 |
| 8 | PL 1 env2 | 28.60 | 400 | 110.17 | 200 | 51.92 | 1.8 | 48.2 |
| 9 | PL 2 env1 | 1.59 | 400 | 6.12 | 200 | 51.92 | 32.7 | 17.3 |
| 10 | PL 2 env2 | 2.05 | 400 | 7.9 | 200 | 51.92 | 25.3 | 24.7 |

**S4 Table:** **AMPure bead clean up and DNA quantification data of pool1 & pool2 ARTIC PCR products in positive sewage samples before indexing**

| S.N<br>. | SAMPLE<br>ID | VOLUM<br>E | AMPURE<br>BEAD | READING<br>1 | READING<br>2 | READING<br>3 | Averag<br>e | ng for 100 fm w/<br>400bp | Vol. of<br>DNA |
| --- | --- | --- | --- | --- | --- | --- | --- | --- | --- |
| 1. | GW11 (P1) | 19 | 26.6 | 13.2 | 13.3 | 13.3 | 13.27 | 25.96 | 2.0 |
| 2. | SA2 (P1) | 19 | 26.6 | 5.48 | 5.48 | 5.5 | 5.49 | 25.96 | 4.7 |
| 3. | SA7 (P1) | 19 | 26.6 | 8.96 | 8.96 | 9 | 8.97 | 25.96 | 2.9 |
| 4. | CH4 (P1) | 38 | 53.2 | 2.02 | 2.02 | 2.02 | 2.02 | 25.96 | 12.9 |
| 5. | KU1 (P1) | 19 | 26.6 | 6.08 | 6.08 | 6.08 | 6.08 | 25.96 | 4.3 |
| 6. | SA5 (P1) | 19 | 26.6 | 8.1 | 8.1 | 8.12 | 8.11 | 25.96 | 3.2 |
| 7. | GW11 (P2) | 19 | 26.6 | 10.1 | 10.1 | 10.1 | 10.10 | 25.96 | 2.6 |
| 8. | SA2 (P2) | 19 | 26.6 | 6.58 | 6.6 | 6.6 | 6.59 | 25.96 | 3.9 |
| 9. | SA7 (P2) | 19 | 26.6 | 3.98 | 4 | 4 | 3.99 | 25.96 | 6.5 |
| 10. | CH4 (P2) | 38 | 53.2 | 8.9 | 8.92 | 8.94 | 8.92 | 25.96 | 2.9 |
| 11. | KU1 (P2) | 19 | 26.6 | 6.72 | 6.72 | 6.74 | 6.73 | 25.96 | 3.9 |
| 12. | SA5 (P2) | 19 | 26.6 | 8.1 | 8.1 | 8.1 | 8.10 | 25.96 | 3.2 |
| 13. | GW10(P1) | 20 | 28 | 1.02 | 1.03 | 1.04 | 1.03 | 25.96 | 25.0 |
| 14. | GW13(P1) | 20 | 28 | 1.3 | 1.3 | 1.31 | 1.30 | 25.96 | 19.8 |
| 15. | GW14(P1) | 20 | 28 | 16.2 | 16.3 | 16.3 | 16.26 | 25.96 | 1.6 |
| 16. | SA1(P1) | 20 | 28 | 0.474 | 0.46 | 0.452 | 0.46 | 25.96 | 57.4 |
| 17. | SA6(P1) | 20 | 28 | 4.16 | 4.18 | 4.2 | 4.18 | 25.96 | 6.2 |
| 18. | SA4(P1) | 20 | 28 | 9.9 | 9.92 | 9.94 | 9.92 | 25.96 | 2.6 |
| 19. | SA3(P1) | 20 | 28 | 0.448 | 0.45 | 0.452 | 0.45 | 25.96 | 57.4 |
| 20. | SA9(P1) | 20 | 28 | 3.9 | 3.9 | 3.92 | 3.91 | 25.96 | 6.6 |
| 21. | GW12(P1) | 20 | 28 | 3.5 | 3.5 | 3.52 | 3.51 | 25.96 | 7.4 |
| 22. | GW15(P1) | 20 | 28 | 36 | 36 | 36 | 36 | 25.96 | 0.7 |
| 23. | GW10(P2) | 20 | 28 | 1.06 | 1.07 | 1.07 | 1.06 | 25.96 | 24.3 |
| 24. | GW13(P2) | 20 | 28 | 4.14 | 4.14 | 4.14 | 4.14 | 25.96 | 6.3 |
| 25. | GW14(P2) | 20 | 28 | 24.6 | 24.6 | 24.6 | 24.6 | 25.96 | 1.1 |
| 26. | SA1(P2) | 20 | 28 | 0.936 | 0.936 | 0.938 | 0.94 | 25.96 | 27.7 |
| 27. | SA6(P2) | 20 | 28 | 3.6 | 3.2 | 2.92 | 3.24 | 25.96 | 8.9 |
| 28. | SA4(P2) | 20 | 28 | 11.9 | 11.9 | 11.6 | 11.8 | 25.96 | 2.2 |
| 29. | SA3(P2) | 20 | 28 | 6.28 | 6.28 | 6.28 | 6.28 | 25.96 | 4.1 |
| 30. | SA9(P2) | 20 | 28 | 4.22 | 4.22 | 4.24 | 4.22 | 25.96 | 6.1 |
| 31. | GW12(P2) | 20 | 28 | 8.24 | 8.24 | 8.26 | 8.24 | 25.96 | 3.1 |
| 32. | GW15(P2) | 20 | 28 | 4.36 | 4.36 | 4.36 | 4.36 | 25.96 | 6.0 |

## Data Availability Statement

The data used to support the findings of this study are included within the article. Accession numbers mentioned in this study are-MW739929 & MW739930.

## Funding

This research project was funded by BIOVAC Nepal and Intrepid Nepal Pvt. Limited. With partial funding support from the PREDICT project, Australian Development Agency and PSI grant from the Government of the Netherlands.

## Competing interests

The authors have declared that no competing interests exist.

## Notes

### Competing Interest Statement

The authors have declared no competing interest.

### Funding Statement

no specific funding for this research.

### Author Declarations

This study is based on environmental samples, hence no specific IRB is required.

